# Architectures of epidemic models: accommodating constraints from empirical and clinical data

**DOI:** 10.1101/2020.12.15.20248249

**Authors:** G. Turinici

## Abstract

Deterministic compartmental models have been used extensively in modeling epidemic propagation. These models are required to fit available data and numerical procedures are often implemented to this end. But not every model architecture is able to fit the data because the structure of the model imposes hard constraints on the solutions. We investigate in this work two such situations: first the distribution of transition times from a compartment to another may impose a variable number of intermediary states; secondly, a non-linear relationship between time-dependent measures of compartments sizes may indicate the need for structurations (i.e., considering several groups of individuals of heterogeneous characteristics).

## 1 Introduction

The recent COVID-19 epidemic stimulated to a large extent the research into the domain of mathematical epidemiology, prompted for instance by the hope to obtain faithful predictions of the ongoing pandemic unfold and assess pharmaceutical and non-pharmaceutical epidemic control interventions.

One of the most easy to use frameworks is that of the deterministic compartmental models [1, 2, 3, 4, 5] (originating from the well known SIR model [6] of Kermack and McKendrick) that partitions all individuals in a population into several classes and prescribes a set of differential equations to model the evolution of the number of individuals in each class; in this set we can include the SIR model and its variants like SEIR, *SI*^*n*^*R*, etc.) or more involved proposals [7, 8, 9, 10, 11, 12, 13].

In order to be effective, these models, that involve multiple parameters, have to be calibrated to available data, that is, to find the precise values of the parameters such that the model fits the observed data. Sometimes the available data can be in contradiction with the model and thus the model has to be dropped or evolved.

The goal of this paper is to discuss how can one design the compartmental model in order to have the right architecture and thus to eventually have a good calibration quality.

We explore two situation: first for the case of *S*(*E*)*I*^*n*^*R* models where a trick [14] can help deal with non-exponential passage distributions and secondly the situation of models where time-dependent measures of class sizes are available.

## 2 Basic model descriptions and notations

We consider here the situation of a SEIR (*Susceptible, Exposed, Infected, Recovered*) model [4] that is illustrated in figure 1 and whose mathematical transcription is:

**Figure 1:**
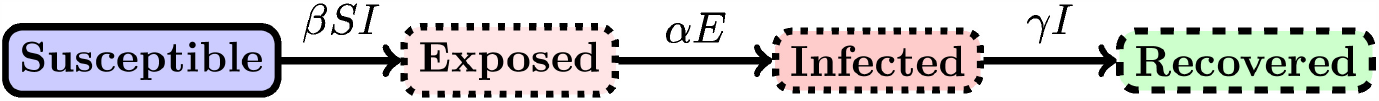
Schematic evolution in the SEIR model.

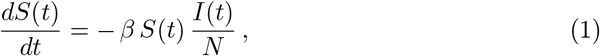

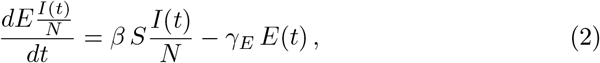

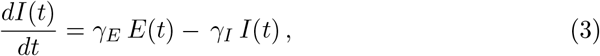

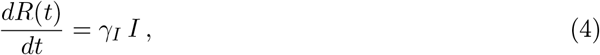

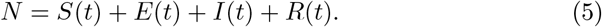

The SEIR model is relevant and has been used in many practical situations; it can also be seen as the average dynamics of individuals that follow a Markov chain dynamics between states *Susceptible, Exposed, Infected*, and *Recovered*. For instance, to first order in Δ*t*, the probability to go from “Susceptible” to “Exposed” during [*t, t* + Δ*t*] is 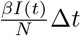, from “Exposed” to “Infected” is *α*Δ*t* and from “Infected” to “Recovered” is *γ*Δ*t*.

### Remark 1

*Similar equations arise when modelling the intra-host infection dynamics, see [15, 16] for a classical introduction and [13] for use in the context of COVID-19. All results of this paper apply with straightforward adaptations*.

## 3 Non-exponential passage times between compartments: need for intermediary states

Let us focus on the “Infected” → “Recovered” transition. The time that an individual spends in the class *I* is an exponential random variable of parameter *γ*; such a distribution can be empirically observed and unfortunately it happens often that the observed distribution is not exponential but rather more close to a gamma distribution (see [17, 18] for COVID-19 related information). In order to respect this constraint, proposals have been documented in the literature (see [14] for an entry point to this literature) that employ the *linear chain trick*; it consists in replacing a single infectious stage with *K* identical exponentially distributed sub-stages, each having a mean period 1*/*(*Kγ*). The historical approach is to divide the infected population into *K* identical stages *I*^1^, *I*^2^, …, *I*^*K*^ to create the so-called *SI*^*K*^*R* model and we denote the total infected populationby 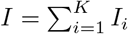.

We depart from this setting by considering a more general model with *K* stages that have each a different parameter *γ*_*k*_.

In this section we ask the following question: can any continuous distribution on [0,*∞* [be represented using the linear chain trick ? Or, put it otherwise, are there any constraints on some observed distribution of residence times in the *Infected* state that is the sum of *K* independent exponential random variables of parameters *γ*_1_, …, *γ*_*K*_ ? Note that such a distribution is called *hypoexponential distribution* and has already been used in mathematical biology, see [19, 20].

The continuous-time transcription of the model’s equations is:

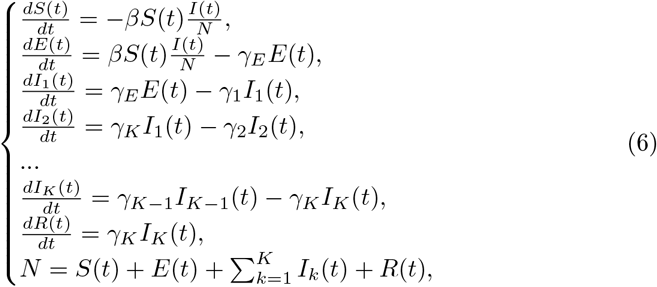

here *S, E, I* and *R* are the numbers of individuals in the *Susceptible, Exposed, Infected* and respectively *Recovered* classes. The initial distribution of individuals at time 0 is (*S*(0), *E*(0), *I*_1_(0), …, *I*_*K*_(0), *R*(0)), which is supposed to be known (otherwise is numerically fitted to data). Note that the system (6) possesses a conservation law, i.e. *S*(*t*) + *E*(*t*) + Σ_*k*_ *I*_*k*_(*t*) + *R*(*t*) = constant, for all *t* ≥0.

Here 1*/γ*_*k*_ is the average number of days a patient spends in class *I*_*k*_. We prove the following

### **Proposition 2**.

*1. Let X be a positive random variable. Then if*

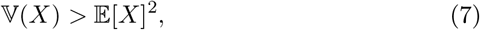

*then X is not a hypo-exponential variable, i*.*e*., *cannot be represented as a sum of independent exponentially distributed random variables X*_*k*_:

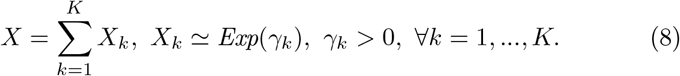

2. *On the contrary, given two strictly positive numbers e, v >* 0 *with v* ≤ *e*^2^ *there exist K and a hypo-exponential distribution satisfying* (8) *such that* 𝔼 [*X*] = *e and* 𝕧 (*X*) = *v*.

3. *If X is a hypoexponential variable that can be represented as in equation* (8) *then* 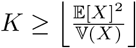

*Here* ⌈· ⌉*is the operator returning the largest integer smaller than the argument, and the operators* 𝔼 [·] *and* 𝕧 [·] *are the mean and variance with respect to the law of X*.

*Proof*. Suppose (8) is true. Then 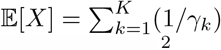 and 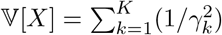 Of course, since 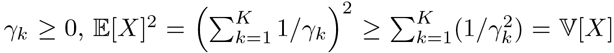 which proves the point 1. On the other hand, using the Cauchy-Schwartz inequality 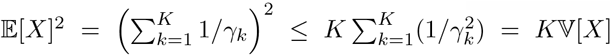 which shows that *K* ≥ 𝔼 [*X*]^2^*/*𝕧 [*X*] and proves the point 3 of the proposition. Denote now 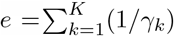 and let us enquire about the possible values of 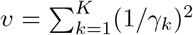 the previous considerations show that*v* ∈ [*e*^2^*/K, e*^2^]. The lowest bound is attained exactly when *γ*_*k*_ are all equal; the upper bound is attained asymptotically when all *γ*_*k*_ go to zero except one of them that tends to 1*/e* (and it is obtained exactly when *K* = 1). Taking any continuous deformation of *γ*_*k*_ (as a *K*-tuple) between these extreme values (while keeping *e* constant) one can show that all values in the interval *v* ∈ [*e*^2^*/K, e*^2^[are attained. Making now *K* → ∞and adding the situation *K* = 1 we obtain that all *v*∈]0, *e*^2^] are attained, hence the item 2 of the proposition is also proved.

### **Remark 3**.

*The point 3 informs on the minimal number of stages that are necessary to implement a given distribution; this is not related to the uniqueness (in many situations a given hypo-exponential distributed random variable can only be decomposed in an unique way as sum of independent exponential variables) but rather as an indication that more the distribution departs from the exponential situation (which corresponds to* 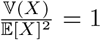*) more stages are necessary to describe it. Reciprocally, the item 2 is not an existence result but only informing that comparing then mean and variance cannot bring additional information on whether some distribution can be represented as hypo-exponential or not. One may can investigate in the same vein the domain of possible values for the triple (mean, variance, skewness) (skewness has an explicit simple form* 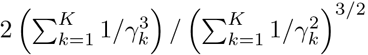

## 4 Nonlinear relationships between sizes of the compartments: the need for structuration

Let us now consider a different view of the passage from one state to another. We will take as model the passage from *I* to *R* in figure 1 although other situations are also relevant (we will give a different example below).

The discussion in this section is independent of that in section 3. If the (4) holds true with a constant *γ*_*I*_ then of course, 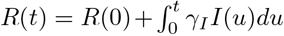. In particular, considering *R*(0) = 0 (otherwise the initial value has to be subtracted), the functions *R*(·) and 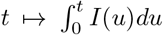 will be linearly dependent; supposing empirical data on those two functions is available (usually the public statistics communicate such information), this requirement can be checked irrespective of that fact that the constant *γ*_*I*_ is known or not (i.e., before any numerical fitting procedure).

We will take a general notation to designate as *A* the source state and *B* the secondary state, i.e. the flow of individuals is from the state “A” to state “B” at rate *γ*, i.e., on average, an individual stays in state “A” for 1*/γ* days. As an example “A” can be the the hospitalized state and ‘B’ the deceased (see next section). The mathematical formula relating the two is

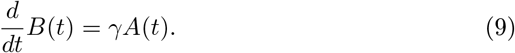

### 4.1 Illustration

As an illustration let us consider the situation for COVID-19 where two figures are most often recovered in the public health authorities statistics, namely the number of hospitalizations and the death toll. These figures are indeed of higher statistical quality than the number of infections which is depending on the definition and of further considerations (e.g., accounting of asymptomatics, etc.). We will consider moreover a model where hospitalization is a state of the Markov chain (sometimes labeled “severe cases”) linked directly to “deaths” state (counting deceased).

The data is plotted in figure 2. Although both curves are related, there is no linear relationship between them and the differences are both qualitative and quantitative.

**Figure 2:**
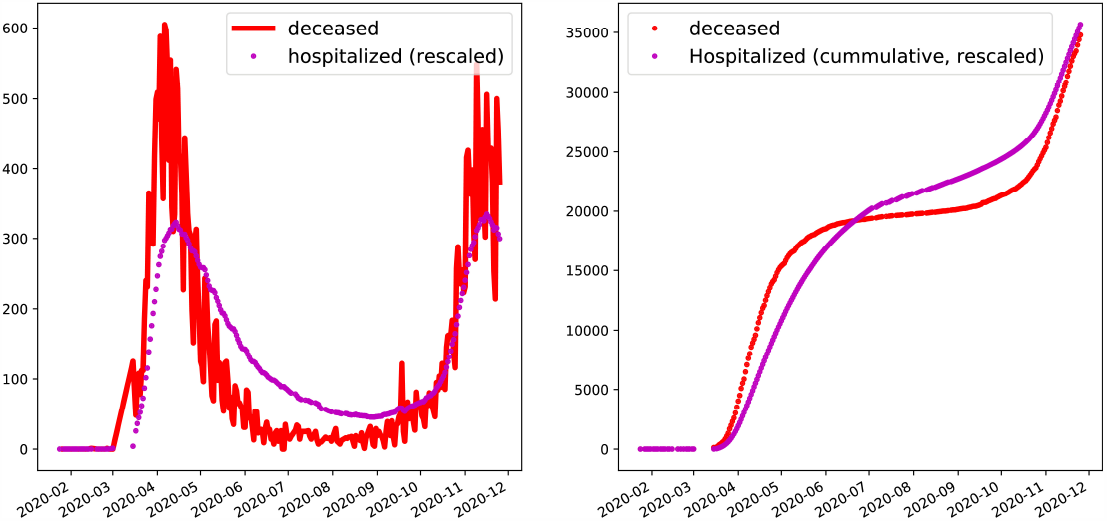
Empirical data for France concerning the COVID-19 hospitalizations and deceased (source “Santé Publique France”). We use the notations “A” = hospitalized, “B”= deceased. **Left:** plot of the numbers of hospitalized (rescaled arbitrarily for graphical convenience) (i.e., *A*(*t*)) and the daily deaths (i.e. with our notations *B*(*t* + 1) − *B*(*t*) as an approximation of 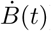 **Right**. Cumulative versions of the above, i.e., 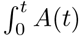 and *B*(*t*). In both cases, no linear relationship between the two exists.

### 4.2 Structuring the model: theoretical constraints

The fact that the linear relationship expected from equation (9) is not true in practice means that, in full rigor, the model cannot be used. It has to be fixed. One way to address this issue is to *structure* the model i.e., to recognize that not all individuals are alike and the average time of 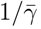 days spent in state ‘A’ does not mean everybody spends exactly this amount of time in that compartment.

We consider then a structuration by this *γ* parameter, that is, we consider that each individual has its own *γ* belonging to some set of possible values *{γ*_1_, …, *γ*_*L*_ *}*.

This view can readily be extended to accommodate an infinite set of possible values [0,*∞* [and a probability measure defined on it. The equation (9) is to be replaced, with obvious notations, by:

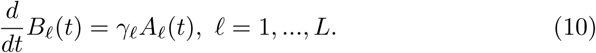

What is less obvious is to understand to what correspond the empirical measured values; for instance, the number of daily hospitalisations would correspond to 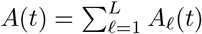, the cumulative death toll to 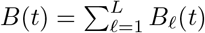; however the distribution of values of *γ* that is obtained is not the real *γ* distribution but instead is the distribution conditioned on the fact that the final state at final time *T* is “B”. If we denote by *X*(*t*) the state of an individual at time *t*, for instance the empirical variance of *γ* (that can be read from the empirical clinical data) will be 𝕧 [*γ* |*X*(*T*) = “*B*”]^1^.

Accordingly, when we refer to mean or variance of 1*/γ* we will in fact refer

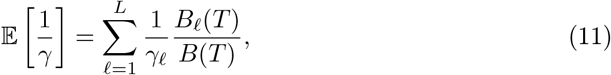

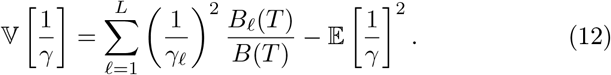

As in section 3 we will prove a result involving those three measurable quantities: the time dependent data *A*(*t*) and *B*(*t*) and the conditional variance of 1*/γ*.

#### **Proposition 4**.

*With the notations in* (12) *we have:*

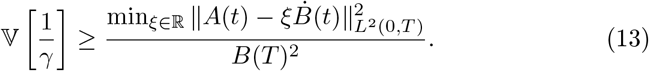

*Proof*. We will replace in 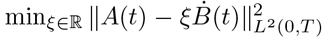 the minimum value by the average 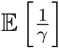 and write:

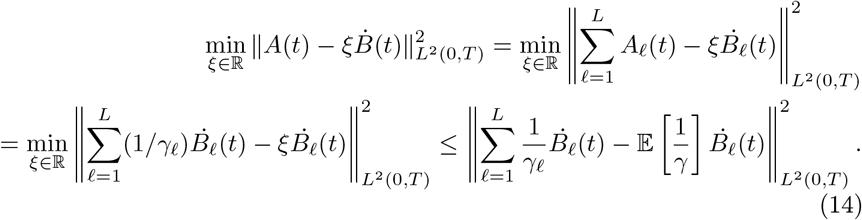

We continue with a upper bound:

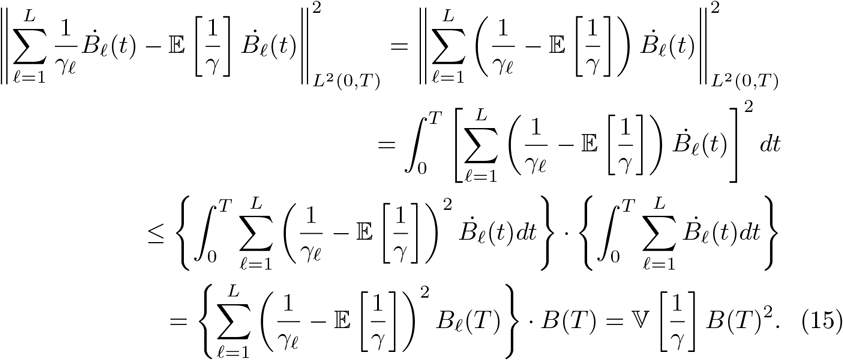

#### **Remark 5**.

*The presence of the term* 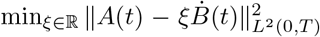 *in equation* (13) *is to measure the difference with the un-structured situation (when it is null). In particular, when the term is non-null this means that no unstructured model can fit both observed data A*(*t*) *and B*(*t*). *The distribution of γ has to satisfy* (13). *This can be used to orient numerical procedures to fit the γ parameters*.

## 5 Concluding remarks

We presented in this contribution how constraints coming from the empirical available data, (that the epidemic models are fitted to reproduce before any further, e.g., previsional, use) impose constraints on the model architecture: in the first case, in order to accommodate non-exponential passage times, the number of compartments needs to be increased and in the second case structuration by some transmission parameters is necessary when there is no linear-relationship between some compartment related time-dependent functions. Additional constraints of this type can be found along the lines proposed in this paper but are left for future work. These considerations can help propose relevant models and are useful in the implementation of numerical procedures to find the model parameters and will ultimately impact the model quality.

## Data Availability

not relevant for content

## Footnotes

1 With a slight abuse of language we identify the random variable 𝕧 [*γ*|*X*(*T*) = “*B*”] with its value on the set *{X*(*T*) = “*B*”*}*. to:

